# Inhaled corticosteroid use and risk COVID-19 related death among 966,461 patients with COPD or asthma: an OpenSAFELY analysis

**DOI:** 10.1101/2020.06.19.20135491

**Authors:** The OpenSAFELY Collaborative, Anna Schultze, Alex J Walker, Brian MacKenna, Caroline E Morton, Krishnan Bhaskaran, Jeremy P Brown, Christopher T Rentsch, Elizabeth Williamson, Henry Drysdale, Richard Croker, Seb Bacon, William Hulme, Chris Bates, Helen J Curtis, Amir Mehrkar, David Evans, Peter Inglesby, Jonathan Cockburn, Helen I McDonald, Laurie Tomlinson, Rohini Mathur, Kevin Wing, Angel YS Wong, Harriet Forbes, John Parry, Frank Hester, Sam Harper, Stephen JW Evans, Jennifer Quint, Liam Smeeth, Ian J Douglas, Ben Goldacre

**Affiliations:** The DataLab, Nuffield Department of Primary Care Health Sciences, University of Oxford, OX26GG; London School of Hygiene and Tropical Medicine, Keppel Street, London WC1E 7HT; TPP, TPP House, 129 Low Lane, Horsforth, Leeds, LS18 5PX; NIHR Health Protection Research Unit (HPRU) in Immunisation; National Heart and Lung Institute, Imperial College, London, SW7 2BU

**Keywords:** COVID-19, Pulmonary Disease, Chronic Obstructive, Asthma, Steroids, Pharmacoepidemiology

## Abstract

**Background:** Early descriptions of the coronavirus outbreak showed a lower prevalence of asthma and COPD than was expected for people diagnosed with COVID-19, leading to speculation that inhaled corticosteroids (ICS) may protect against infection with SARS-CoV-2, and development of serious sequelae. We evaluated the association between ICS and COVID-19 related death using linked electronic health records in the UK.

**Methods:** We conducted cohort studies on two groups of people (COPD and asthma) using the OpenSAFELY platform to analyse data from primary care practices linked to national death registrations. People receiving an ICS were compared to those receiving alternative respiratory medications. Our primary outcome was COVID-19 related death.

**Findings:** We identified 148,588 people with COPD and 817,973 people with asthma receiving relevant respiratory medications in the four months prior to 01 March 2020. People with COPD receiving ICS were at a greater risk of COVID-19 related death compared to those receiving a long-acting beta agonist (LABA) and a long-acting muscarinic antagonist (LAMA) (adjusted HR = 1.38, 95% CI = 1.08 – 1.75). People with asthma receiving high dose ICS were at an increased risk of death compared to those receiving a short-acting beta agonist (SABA) only (adjusted HR = 1.52, 95%CI = 1.08 – 2.14); the adjusted HR for those receiving low-medium dose ICS was 1.10 (95% CI = 0.82 – 1.49). Quantitative bias analyses indicated that an unmeasured confounder of only moderate strength of association with exposure and outcome could explain the observed associations in both populations.

**Interpretation:** These results do not support a major role of ICS in protecting against COVID-19 related deaths. Observed increased risks of COVID-19 related death among people with COPD and asthma receiving ICS can be plausibly explained by unmeasured confounding due to disease severity.

**Funding:** This work was supported by the Medical Research Council MR/V015737/1.

## Introduction

The ongoing pandemic due to the novel coronavirus, SARS-CoV-2, has now affected over 7 million people worldwide with at least 400,000 people having died with COVID-19^1^. People with more severe COVID-19 outcomes, including hospitalisation or death, tend to be older and have pre-existing comorbidities ^2,3,4,5,6,7,8^. Severe outcomes are often a result of lung complications, such as acute respiratory distress syndrome (ARDS) and respiratory failure. However, early reports of COVID-19 patients described an unexpectedly low prevalence of chronic respiratory conditions among hospitalised patients^9^. Although other studies suggest that chronic lung diseases, including chronic obstructive pulmonary disease (COPD), increase the risk of severe outcomes^6–8^, reported effect sizes for asthma have been modest ^6,7^. This has led to speculation that treatments for respiratory disease, specifically inhaled corticosteroids (ICS), may have a protective effect against SARS-CoV-2.^9–11^.

ICS are used to reduce airway inflammation, oedema, and mucus secretions. In-vitro evidence indicates that the ICS ciclesonide can suppress SARS-CoV-2 replication^12^, and budesonide combined with glycopyrronium and formoterol inhibits the production of cytokines in cells exposed to HCoV-229E, another human coronavirus^13^. The oral/IV steroid dexamethasone has recently been shown to reduce the risk of death in severe COVID-19^14^. Conversely, although ICS have low systemic absorption, they have been associated with increased risk of developing pneumonia in people with COPD ^15–17^, as well as other systemic steroid-related adverse effects^18^. A recent systematic review of the role of ICS in SARS-CoV-2, SARS-CoV-1, and MERS found no studies investigating the impact of prior ICS use on outcomes in any of these infections ^10^.

We therefore set out to explore the association between current ICS use and outcomes in COVID-19, using the OpenSAFELY platform which contains linked primary care electronic health record data for approximately 40% of the population in England.

## Methods

### Study Design

We conducted two cohort studies using primary care electronic health record (EHR) data linked to death data from the Office for National Statistics (ONS). The index date (start of follow up) for both cohorts was 01 Mar 2020; follow-up lasted until 06 May 2020.

### Data Source

Primary care records managed by the GP software provider The Phoenix Partnership (TPP) were linked to ONS death data through OpenSAFELY, a data analytics platform created by our team on behalf of NHS England^19^ to address urgent COVID-19 research questions ^7^ (https://opensafely.org). OpenSAFELY provides a secure software interface allowing the analysis of pseudonymised primary care patient records from England in near real-time within the EHR vendor’s highly secure data centre, avoiding the need for large volumes of potentially disclosive pseudonymised patient data to be transferred off-site. This, in addition to other technical and organisational controls, minimises the risk of re-identification. Similarly pseudonymised datasets from other data providers are securely provided to the EHR vendor and linked to the primary care data. The dataset analysed within OpenSAFELY is based on 24 million people currently registered with GP surgeries using TPP SystmOne software. It includes pseudonymised data such as coded diagnoses, medications and physiological parameters. No free text data is included.

### Study Populations

The COPD cohort included adults older than 35 years with COPD and current or former smoking recorded any time before the index date^20^. We excluded people with prior diagnoses of other chronic respiratory conditions, or with asthma in the three years before the index date^21^, and those receiving nebulised COPD medications in the twelve months before the index date or a leukotriene receptor antagonist (indicating potential asthma) in the four months before the index date.

The asthma cohort included adults older than 18 years with asthma recorded within three years prior to the index date. People with COPD or other chronic respiratory conditions prior to the index date were excluded, as were those receiving a LAMA without an ICS, as this indicates possible COPD^22^.

People with missing data for gender, index of multiple deprivation (IMD), or less than one year of primary care records were excluded (supplemental figure 1).

### Exposures

In the COPD population, people issued at least one ICS prescription within four months prior to the index date either in combination with LABA or LAMA/LABA, or as single therapy provided there was also at least one prescription record of a LABA, were compared with those with a prescription for a LABA/LAMA (combined or as separate single therapy prescriptions) only^22^. We did not include patients receiving LAMA monotherapy, as we were expecting greater clinical comparability between the LAMA/LABA and ICS-based therapy groups.

In the asthma population, people prescribed high dose ICS and low/medium-dose ICS during the four months before index date were compared with those prescribed SABA only. Exposure for people prescribed both high and low/medium dose ICS was assigned according to their most recent prescription. Inhalers were assigned to low/medium or high dose based the OpenPrescribing.net prescribing explorer which was developed based on BTS/SIGN guidance^23^. Studies have shown that a significant percentage of people with asthma receiving SABA only are eligible for ICS treatment^24^, suggesting they have similar disease severity to those receiving ICS and therefore represent a reasonable comparator group. The characteristics of all other people are described in supplementary material (supplemental table 1-2), however they are excluded from regression models to avoid comparisons to individuals not prescribed drugs of interest ^25^.

### Outcomes

The outcome was COVID-19 related death as registered in ONS data using ICD-10 codes U07.1 (“COVID-19, virus identified”) and U07.2 (“COVID-19, virus not identified”) listed either as the underlying or any contributing cause of death. The latter ICD-10 code is used when laboratory testing is inconclusive or unavailable^26^.

### Covariates

Potential determinants of exposures and outcomes were identified by reviewing literature and through discussions with practising clinicians. As this is a study of current users, determinants of exposures include both factors that may affect the initial choice of treatment as well as those that influence whether patients remain on a certain treatment. The final list of potential confounders can be seen in box 1. Our methodology for creating codelists associated with these confounders has been previously described^7^: this included clinical and epidemiological review and sign-off by at least two authors. Detailed information on every codelist is shared at https://codelists.opensafely.org/.

### Statistical Methods

Individuals characteristics were summarised using descriptive statistics, stratified by exposure status. Time to the primary outcome is displayed in Kaplan-Meier (KM) plots with time in study as the timescale. The competing risk of death from non-COVID-19 causes was dealt with by analysing the cause-specific hazard, with people dying from other causes censored at their date of death^27^. We used cause-specific Cox regression models to estimate hazard ratios (HRs) and 95% confidence intervals (CIs) for the association between exposure categories and the outcome in each population. Univariable models, models adjusted for age (using restricted cubic splines) and sex as well as fully adjusted models including all covariates were fitted. Region was included as a stratification variable in fully adjusted models. We evaluated an *a priori* specified interaction between ICS exposure and age, to see if we could distinguish a differential effect in groups known to be at higher risk.

### Sensitivity Analyses

In sensitivity analyses, first, we split the exposure categories in the COPD population to examine the effect of ICS with LABA/LAMA (triple combination) and ICS with LABA (dual combination) separately, anticipating greater underlying disease severity in people receiving triple therapy. Second, we restricted analyses to the largest ethnicity group (i.e., white British) to exclude any substantial confounding by ethnicity. We did not adjust for ethnicity in the main models as this was not anticipated to be a strong confounder and due to a sizable proportion of individuals with missing ethnicity (∼25%). In the asthma population, we varied the sample definition to include people with asthma diagnosed at any time, and a recent prescription for any asthma medication.

### Negative Control Outcomes

We hypothesised that disease severity, but not ICS use, may influence the risk of non-COVID-19 related death. Analyses were therefore conducted using non-COVID-19 death as a negative control outcome censoring people at time of COVID-19 related death. If any potentially harmful association observed in primary analyses was due to confounding (i.e. people prescribed ICS had more severe underlying respiratory disease than those who did not) we expected to observe a similar association with non-COVID-19 related death in people prescribed ICS.

### Quantitative Bias Analysis

We used e-value formulae to calculate the minimum strengths of association between an unmeasured confounder and exposure or outcome, conditional on measured covariates, necessary to fully explain observed associations^28^.

### Software and Reproducibility

Data management was performed using Python 3.8 and SQL, with analysis carried out using Stata 16.1. All of the code used for data management and analyses is openly shared online for review and re-use (https://github.com/opensafely/ics-research). All iterations of the pre-specified study protocol are archived with version control (https://github.com/opensafely/ics-research/tree/master/protocol).

### Patient and Public Involvement

Patients were not formally involved in developing this specific study design that was developed rapidly in the context of a global health emergency. We have developed a publicly available website https://opensafely.org/ through which we invite any patient or member of the public to contact us regarding this study or the broader OpenSAFELY project. The protocol and draft paper have been sent to the Asthma UK and British Lung Foundation Partnership for review from an expert-patient perspective.

## Results

### Patient Characteristics

148,488 people with COPD and 817,973 people with asthma and a relevant prescription within the four months before 01 Mar 2020 were included (supplemental figure 2-3).

#### COPD Population

Table 1 (see end) shows the characteristics of the COPD population. 43,278 (29%) received a LABA/LAMA prescription in the 4 months before index, and 105,210 (71%) prescriptions for ICS+LABA or ICS+LABA/LAMA. Demographic characteristics of treatment groups were similar; median age was 71 (IQR = 63 −77) in the LABA/LAMA group and 72 (IQR = 64 − 78) in the ICS group, and just over half were men (54.5% in the LABA/LAMA group and 53.7% in the ICS group).

The presence of comorbidities was similar across the two treatment groups, except prior asthma diagnosis, which was more common among ICS recipients (12.9% in the LABA/LAMA group compared to 27.7% in the ICS group). The percentage of people with an exacerbation in the last year was lower in the LABA/LAMA group (19.7% vs 26.0%).

**Table 1.** Demographic and Clinical Characteristics - COPD

|  |  | Total | LABA/LAMA Combination | ICS Combination | Other |
| --- | --- | --- | --- | --- | --- |
| Total Included |  | 291,805 (100.0) | 43,278 (100.0) | 105,210 (100.0) | 143,317 (100.0) |
| Demographics |  |  |  |  |  |
| Grouped age | 18 - <40 | 1,232 (0.4) | 85 (0.2) | 184 (0.2) | 963 (0.7) |
|  | 40 - <50 | 10,028 (3.4) | 1,060 (2.4) | 2,289 (2.2) | 6,679 (4.7) |
|  | 50 - < 60 | 40,910 (14.0) | 5,746 (13.3) | 12,237 (11.6) | 22,927 (16.0) |
|  | 60 - <70 | 80,788 (27.7) | 12,599 (29.1) | 29,519 (28.1) | 38,670 (27.0) |
|  | 70 - <80 | 104,319 (35.7) | 16,092 (37.2) | 40,370 (38.4) | 47,857 (33.4) |
|  | 80+ | 54,528 (18.7) | 7,696 (17.8) | 20,611 (19.6) | 26,221 (18.3) |
| Age (years) | Median (IQR) | 71 (63-77) | 71 (63-77) | 72 (64-78) | 70 (61-77) |
|  | Mean (SD) | 69.76 (10.76) | 70.03 (10.10) | 70.76 (10.10) | 68.95 (11.38) |
|  | Min, Max | 35, 103 | 35, 100 | 35, 102 | 35, 103 |
| Male | No | 133,738 (45.8) | 19,699 (45.5) | 48,707 (46.3) | 65,332 (45.6) |
|  | Yes | 158,067 (54.2) | 23,579 (54.5) | 56,503 (53.7) | 77,985 (54.4) |
| Grouped BMI | Underweight (<18.5) | 11,359 (3.9) | 1,682 (3.9) | 4,744 (4.5) | 4,933 (3.4) |
|  | Normal (18.5-24.9) | 89,195 (30.6) | 12,938 (29.9) | 31,858 (30.3) | 44,399 (31.0) |
|  | Overweight (25-29.9) | 95,443 (32.7) | 14,011 (32.4) | 33,494 (31.8) | 47,938 (33.4) |
|  | Obese I (30-34.9) | 55,541 (19.0) | 8,590 (19.8) | 20,278 (19.3) | 26,673 (18.6) |
|  | Obese II (35-39.9) | 22,054 (7.6) | 3,568 (8.2) | 8,379 (8.0) | 10,107 (7.1) |
|  | Obese III (40+) | 10,006 (3.4) | 1,580 (3.7) | 3,945 (3.7) | 4,481 (3.1) |
|  | Missing | 8,207 (2.8) | 909 (2.1) | 2,512 (2.4) | 4,786 (3.3) |
| Smoking status | Never | 0 (0.0) | 0 (0.0) | 0 (0.0) | 0 (0.0) |
|  | Former | 184,400 (63.2) | 26,015 (60.1) | 69,710 (66.3) | 88,675 (61.9) |
|  | Current | 107,405 (36.8) | 17,263 (39.9) | 35,500 (33.7) | 54,642 (38.1) |
|  | Missing | 0 (0.0) | 0 (0.0) | 0 (0.0) | 0 (0.0) |
| Ethnicity | White | 218,717 (75.0) | 32,455 (75.0) | 79,682 (75.7) | 106,580 (74.4) |
|  | Mixed | 676 (0.2) | 92 (0.2) | 186 (0.2) | 398 (0.3) |
|  | Asian or Asian British | 3,366 (1.2) | 258 (0.6) | 835 (0.8) | 2,273 (1.6) |
|  | Black | 1,208 (0.4) | 103 (0.2) | 302 (0.3) | 803 (0.6) |
|  | Other | 1,060 (0.4) | 113 (0.3) | 286 (0.3) | 661 (0.5) |
|  | Unknown | 66,778 (22.9) | 10,257 (23.7) | 23,919 (22.7) | 32,602 (22.7) |
| Index of Multiple Deprivation (IMD) | Least Deprived | 58,902 (20.2) | 8,049 (18.6) | 19,893 (18.9) | 30,960 (21.6) |
|  | 2 | 58,779 (20.1) | 8,419 (19.5) | 20,619 (19.6) | 29,741 (20.8) |
|  | 3 | 59,175 (20.3) | 8,755 (20.2) | 21,235 (20.2) | 29,185 (20.4) |
|  | 4 | 57,633 (19.8) | 8,431 (19.5) | 21,634 (20.6) | 27,568 (19.2) |
|  | Most Deprived | 57,316 (19.6) | 9,624 (22.2) | 21,829 (20.7) | 25,863 (18.0) |
|  | Missing | 0 (0.0) | 0 (0.0) | 0 (0.0) | 0 (0.0) |
| **Treatments** |  |  |  |  |  |
| Single SABA | No | 125,025 (42.8) | 12,349 (28.5) | 21,544 (20.5) | 91,132 (63.6) |
|  | Yes | 166,780 (57.2) | 30,929 (71.5) | 83,666 (79.5) | 52,185 (36.4) |
| High Dose ICS | No | 266,112 (91.2) | 43,278 (100.0) | 79,748 (75.8) | 143,086 (99.8) |
|  | Yes | 25,693 (8.8) | 0 (0.0) | 25,462 (24.2) | 231 (0.2) |
| Low/Medium Dose ICS | No | 204,577 (70.1) | 43,278 (100.0) | 22,739 (21.6) | 138,560 (96.7) |
|  | Yes | 87,228 (29.9) | 0 (0.0) | 82,471 (78.4) | 4,757 (3.3) |
| Single ICS | No | 284,499 (97.5) | 43,278 (100.0) | 102,887 (97.8) | 138,334 (96.5) |
|  | Yes | 7,306 (2.5) | 0 (0.0) | 2,323 (2.2) | 4,983 (3.5) |
| Single SAMA | No | 288,487 (98.9) | 43,120 (99.6) | 103,725 (98.6) | 141,642 (98.8) |
|  | Yes | 3,318 (1.1) | 158 (0.4) | 1,485 (1.4) | 1,675 (1.2) |
| Single LABA | No | 283,744 (97.2) | 40,849 (94.4) | 104,356 (99.2) | 138,539 (96.7) |
|  | Yes | 8,061 (2.8) | 2,429 (5.6) | 854 (0.8) | 4,778 (3.3) |
| Single LAMA | No | 204,171 (70.0) | 38,690 (89.4) | 59,736 (56.8) | 105,745 (73.8) |
|  | Yes | 87,634 (30.0) | 4,588 (10.6) | 45,474 (43.2) | 37,572 (26.2) |
| LABA ICS | No | 216,279 (74.1) | 43,278 (100.0) | 29,684 (28.2) | 143,317 (100.0) |
|  | Yes | 75,526 (25.9) | 0 (0.0) | 75,526 (71.8) | 0 (0.0) |
| LABA LAMA | No | 245,652 (84.2) | 1,929 (4.5) | 100,406 (95.4) | 143,317 (100.0) |
|  | Yes | 46,153 (15.8) | 41,349 (95.5) | 4,804 (4.6) | 0 (0.0) |
| LABA LAMA ICS | No | 258,774 (88.7) | 43,278 (100.0) | 72,179 (68.6) | 143,317 (100.0) |
|  | Yes | 33,031 (11.3) | 0 (0.0) | 33,031 (31.4) | 0 (0.0) |
| Single LTRA | No | 291,805 (100.0) | 43,278 (100.0) | 105,210 (100.0) | 143,317 (100.0) |
|  | Yes | 0 (0.0) | 0 (0.0) | 0 (0.0) | 0 (0.0) |
| **Clinical Conditions** |  |  |  |  |  |
| Chronic kidney disease | No | 241,927 (82.9) | 35,547 (82.1) | 86,857 (82.6) | 119,523 (83.4) |
|  | Yes | 49,878 (17.1) | 7,731 (17.9) | 18,353 (17.4) | 23,794 (16.6) |
| Diagnosed hypertension | No | 146,895 (50.3) | 21,595 (49.9) | 50,961 (48.4) | 74,339 (51.9) |
|  | Yes | 144,910 (49.7) | 21,683 (50.1) | 54,249 (51.6) | 68,978 (48.1) |
| Heart Failure | No | 267,221 (91.6) | 39,424 (91.1) | 95,255 (90.5) | 132,542 (92.5) |
|  | Yes | 24,584 (8.4) | 3,854 (8.9) | 9,955 (9.5) | 10,775 (7.5) |
| Other Heart Diseases | No | 226,356 (77.6) | 33,233 (76.8) | 81,114 (77.1) | 112,009 (78.2) |
|  | Yes | 65,449 (22.4) | 10,045 (23.2) | 24,096 (22.9) | 31,308 (21.8) |
| Cancer | No | 251,789 (86.3) | 37,060 (85.6) | 90,144 (85.7) | 124,585 (86.9) |
|  | Yes | 40,016 (13.7) | 6,218 (14.4) | 15,066 (14.3) | 18,732 (13.1) |
| Diabetes Severity | No Diabetes | 221,197 (75.8) | 32,888 (76.0) | 79,531 (75.6) | 108,778 (75.9) |
|  | Diabetes, not severe | 51,265 (17.6) | 7,583 (17.5) | 19,011 (18.1) | 24,671 (17.2) |
|  | Diabetes, severe | 18,561 (6.4) | 2,711 (6.3) | 6,363 (6.0) | 9,487 (6.6) |
|  | Diabetes, no Hb1AC | 782 (0.3) | 96 (0.2) | 305 (0.3) | 381 (0.3) |
| Recent Statin | No | 150,420 (51.5) | 20,517 (47.4) | 50,894 (48.4) | 79,009 (55.1) |
|  | Yes | 141,385 (48.5) | 22,761 (52.6) | 54,316 (51.6) | 64,308 (44.9) |
| Flu vaccine | No | 73,828 (25.3) | 8,683 (20.1) | 19,904 (18.9) | 45,241 (31.6) |
|  | Yes | 217,977 (74.7) | 34,595 (79.9) | 85,306 (81.1) | 98,076 (68.4) |
| Pneumococcal Vaccine | No | 231,323 (79.3) | 32,266 (74.6) | 83,906 (79.8) | 115,151 (80.3) |
|  | Yes | 60,482 (20.7) | 11,012 (25.4) | 21,304 (20.2) | 28,166 (19.7) |
| Exacerbation in last year | No | 236,123 (80.9) | 34,748 (80.3) | 77,858 (74.0) | 123,517 (86.2) |
|  | Yes | 55,682 (19.1) | 8,530 (19.7) | 27,352 (26.0) | 19,800 (13.8) |
| Asthma ever | No | 236,592 (81.1) | 37,705 (87.1) | 76,033 (72.3) | 122,854 (85.7) |
|  | Yes | 55,213 (18.9) | 5,573 (12.9) | 29,177 (27.7) | 20,463 (14.3) |
| Immunosuppressed (combination algorithm) | No | 290,160 (99.4) | 43,073 (99.5) | 104,672 (99.5) | 142,415 (99.4) |
|  | Yes | 1,645 (0.6) | 205 (0.5) | 538 (0.5) | 902 (0.6) |
| GP consultation count | Median (IQR) | 9 (5-16) | 10 (6-16) | 10 (6-17) | 8 (4-14) |
|  | Mean (SD) | 12.19 (11.62) | 12.72 (11.32) | 13.43 (12.26) | 11.12 (11.12) |
|  | Min, Max | 0, 306 | 0, 276 | 0, 306 | 0, 268 |
| Exacerbation Count | Median (IQR) | 0 (0-0) | 0 (0-0) | 0 (0-1) | 0 (0-0) |
|  | Mean (SD) | .26 (.63) | .26 (.60) | .38 (.77) | .17 (.48) |
|  | Min, Max | 0, 12 | 0, 9 | 0, 12 | 0, 8 |

#### Asthma Population

Table 2 (see end) shows the characteristics of the asthma population. Most (608,583, 74%) had a prescription for low/medium dose ICS in the four months prior to index date; fewer received high dose ICS (101,010, 12%) and SABA only (108,380, 13%).

**Table 2.**
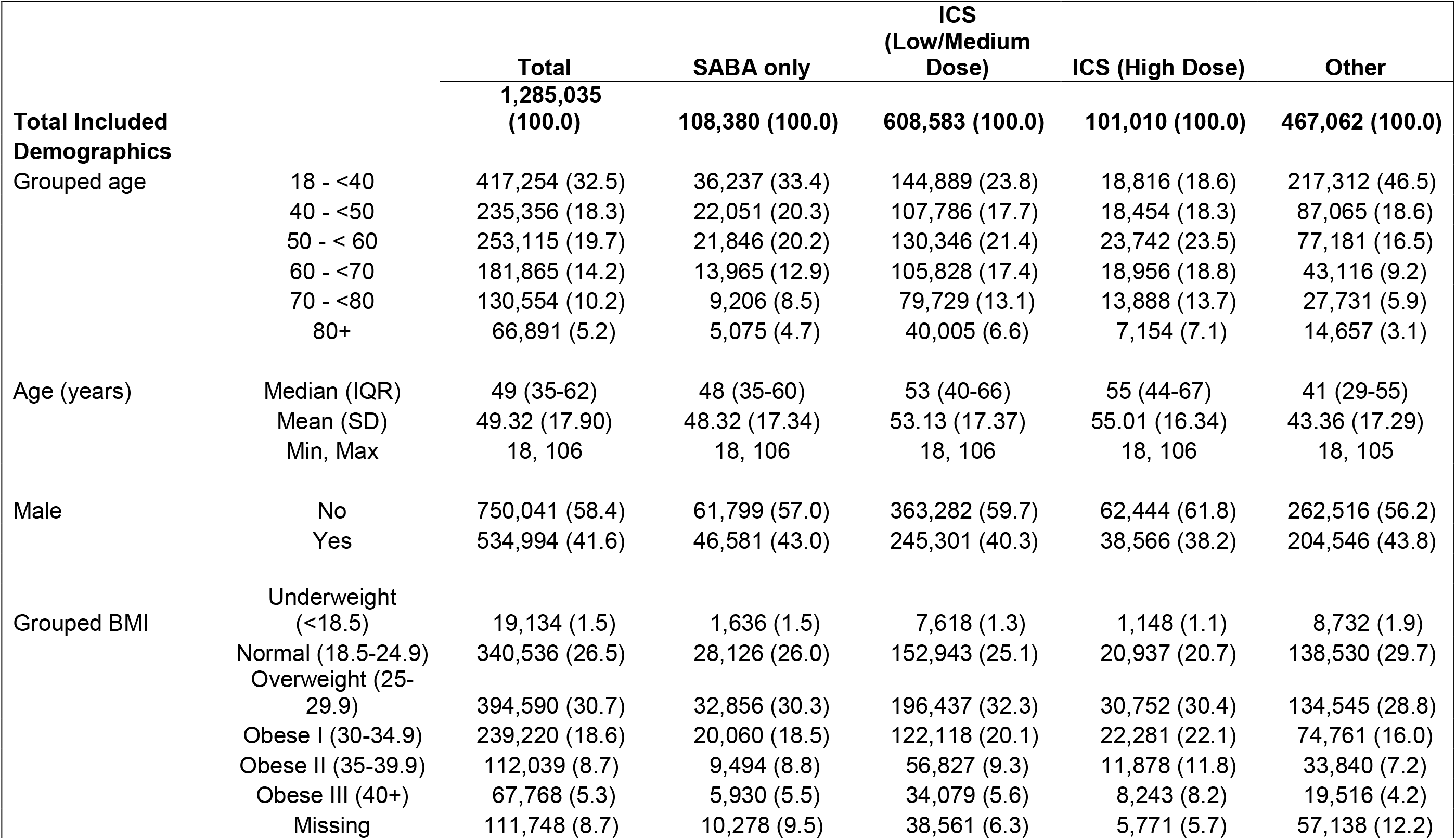
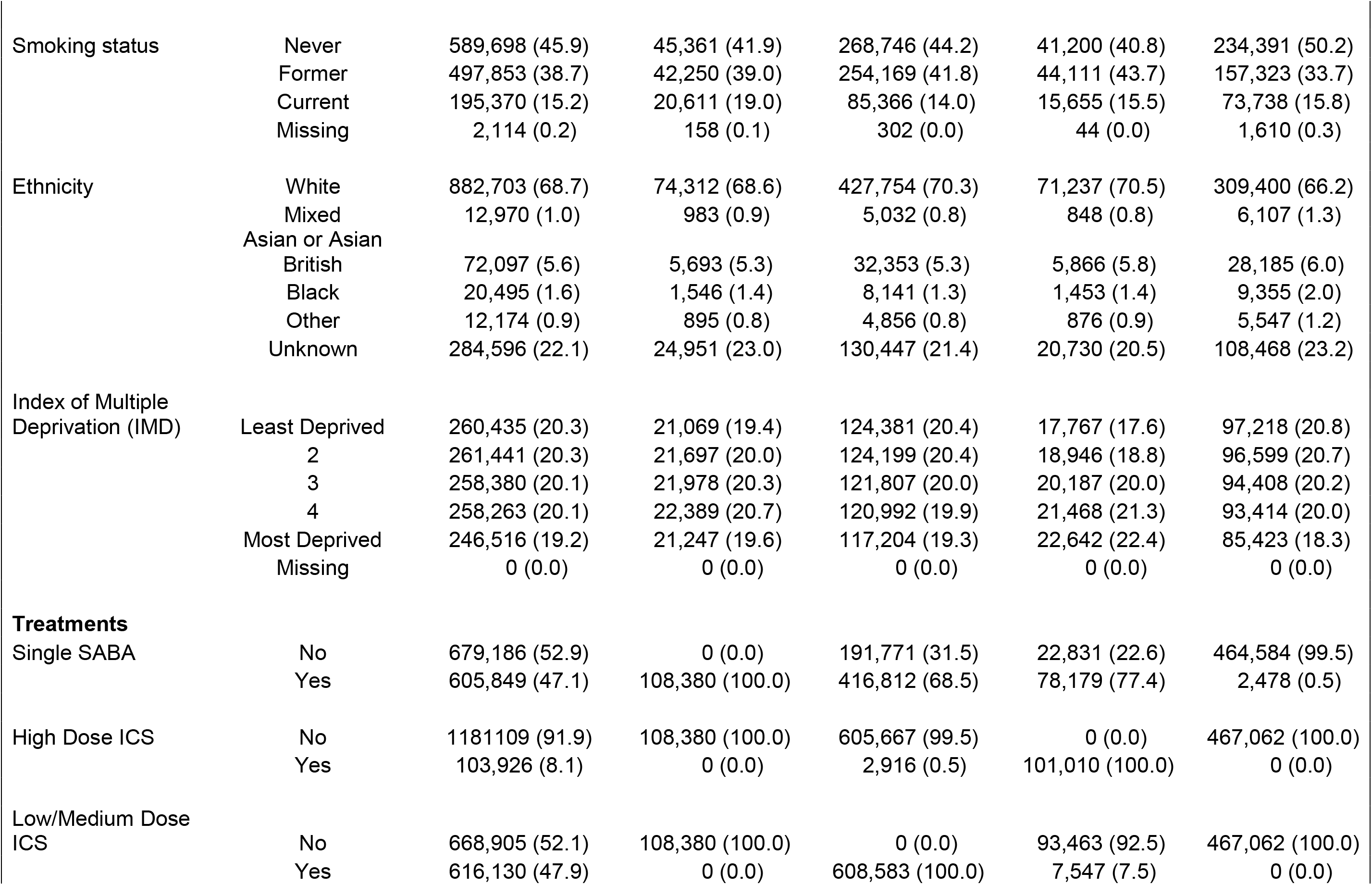
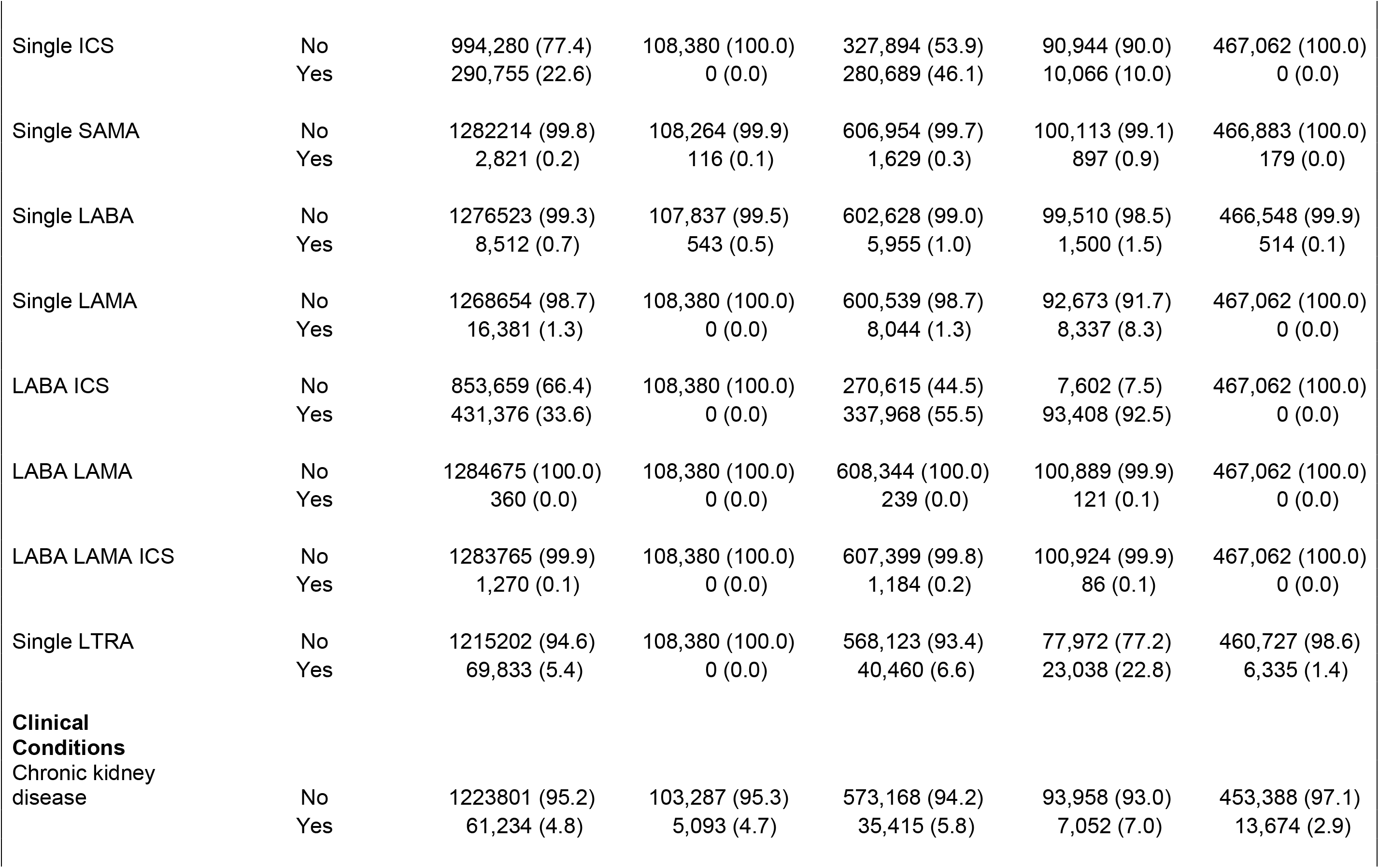
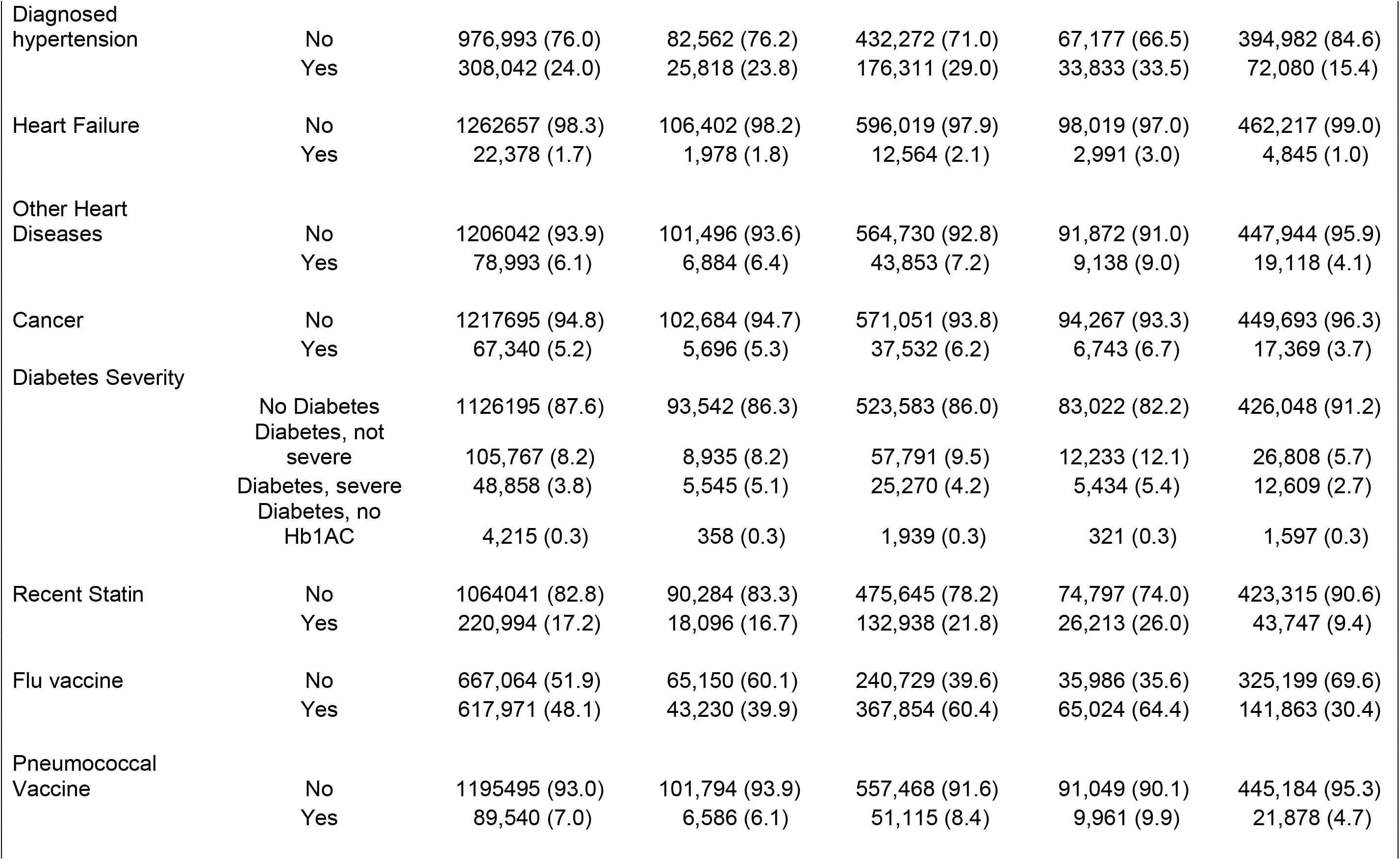
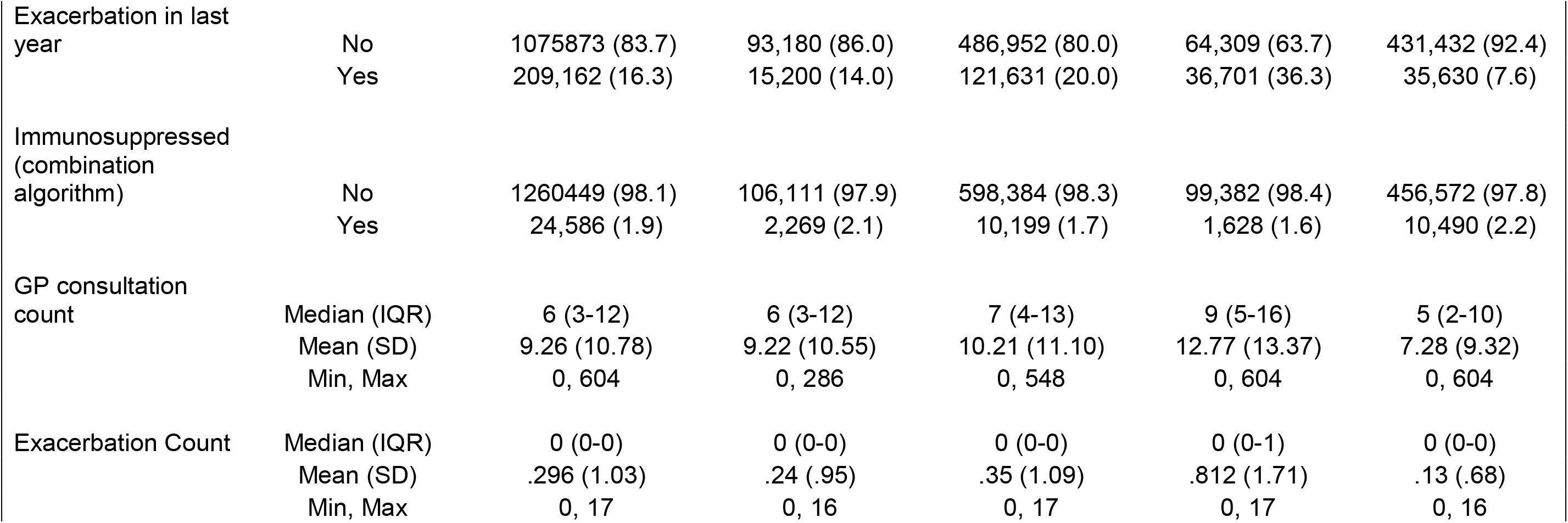
Demographic and Clinical Characteristics - Asthma

The asthma treatment groups differed in terms of demographic and clinical characteristics. The median age was 48 (IQR = 35 - 60), 53 (IQR = 40 - 66), and 55 (IQR = 44 - 67) for SABA only, low/medium dose ICS and high dose ICS respectively. The proportion of men was highest in the SABA group (43.0%), followed by the low/medium dose ICS (40.3%) and high dose ICS (38.2%).

The prevalence of most comorbidities was lowest among people in the SABA only group and highest in the high dose ICS group. The proportion with an asthma exacerbation in the past year increased markedly across treatment groups (14.0%, 20.0% and 36.3% among SABA, low/medium dose and high dose ICS respectively).

### Univariable and Multivariable Results

#### COPD

421 COVID-19 related deaths occurred in the treated COPD population; time to COVID-19 related death by treatment group can be seen in Figure 1a. In univariable models, ICS use was associated with an increased risk of COVID-19 related death (HR = 1.52, 95% CI = 1.20 - 1.91; Figure 2). This association decreased on adjustment for age and gender, and the remaining comorbidities (aHR = 1.38; 95% CI = 1.08 - 1.75; Figure 2). Post-hoc analyses revealed that adjustment for prior exacerbations as a binary variable and smoking status (current/former) had the largest impact on reducing the hazard ratio (supplemental table 6). There was no evidence of a (pre-specified) interaction with age (supplemental table 4). We detected significant deviations from the proportional hazards assumptions by testing for a zero slope in the scaled Schoenfeld residuals as well as by graphical inspection of plots of the Schoenfeld residuals against time (supplemental table 5, Figure 4-6). The KM curve indicated that the hazard ratio was likely above one throughout the follow-up, with the effect size growing over time.

**Figure 1a:**
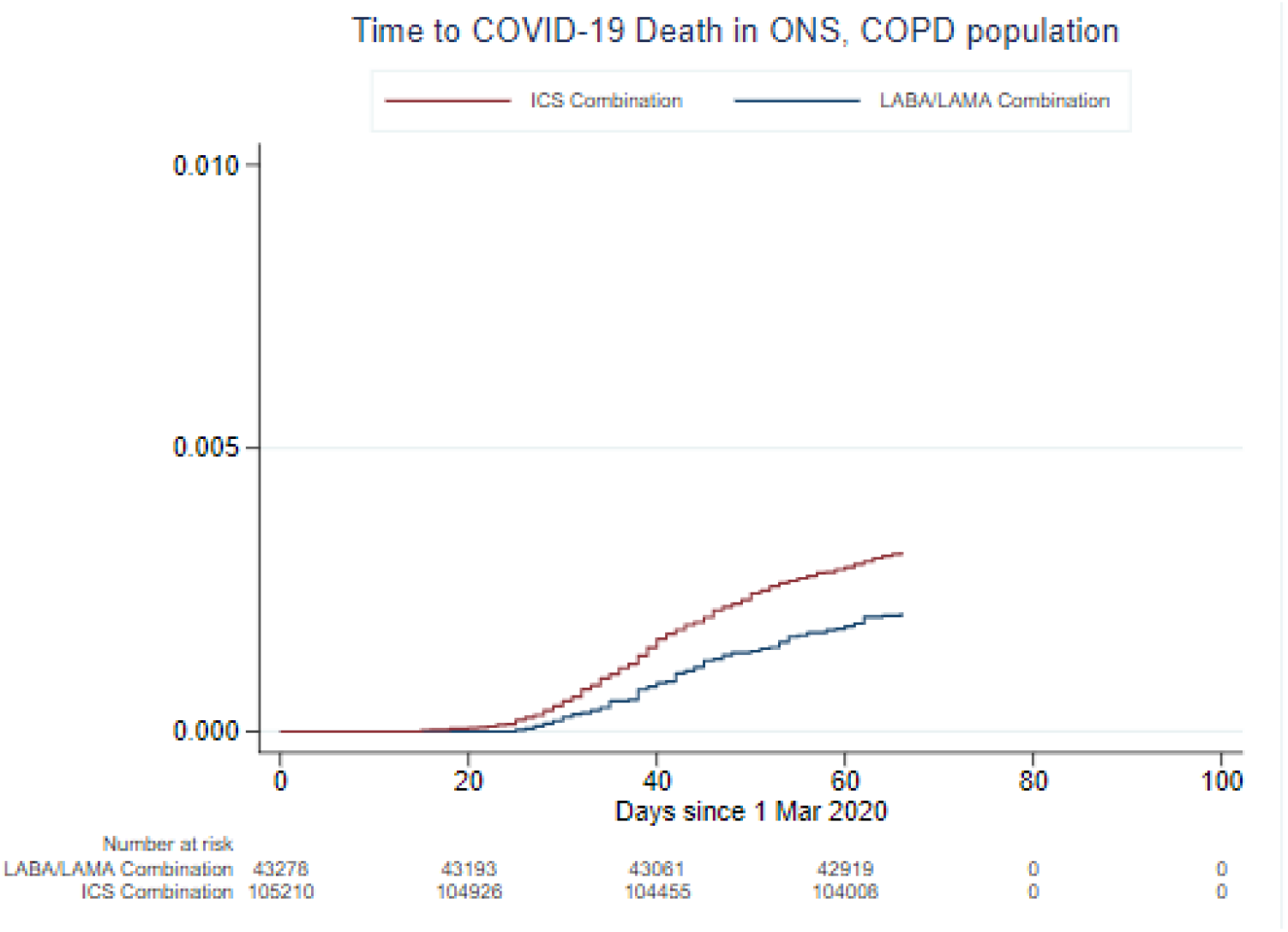
Kaplan-Meier plots in in the COPD population.

**Figure 1b:**
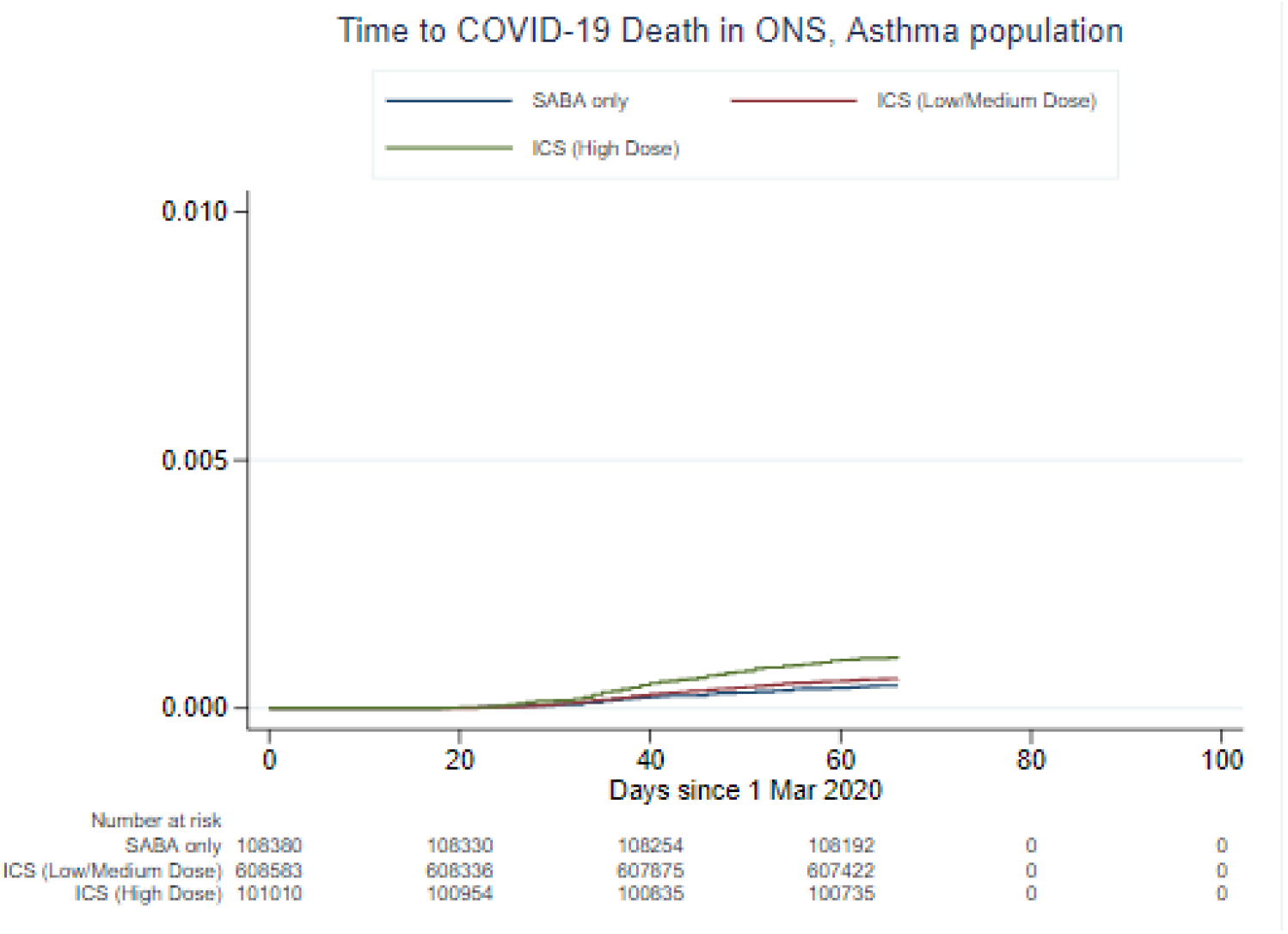
Kaplan-Meier plots in in the asthma population.

**Figure 2:**
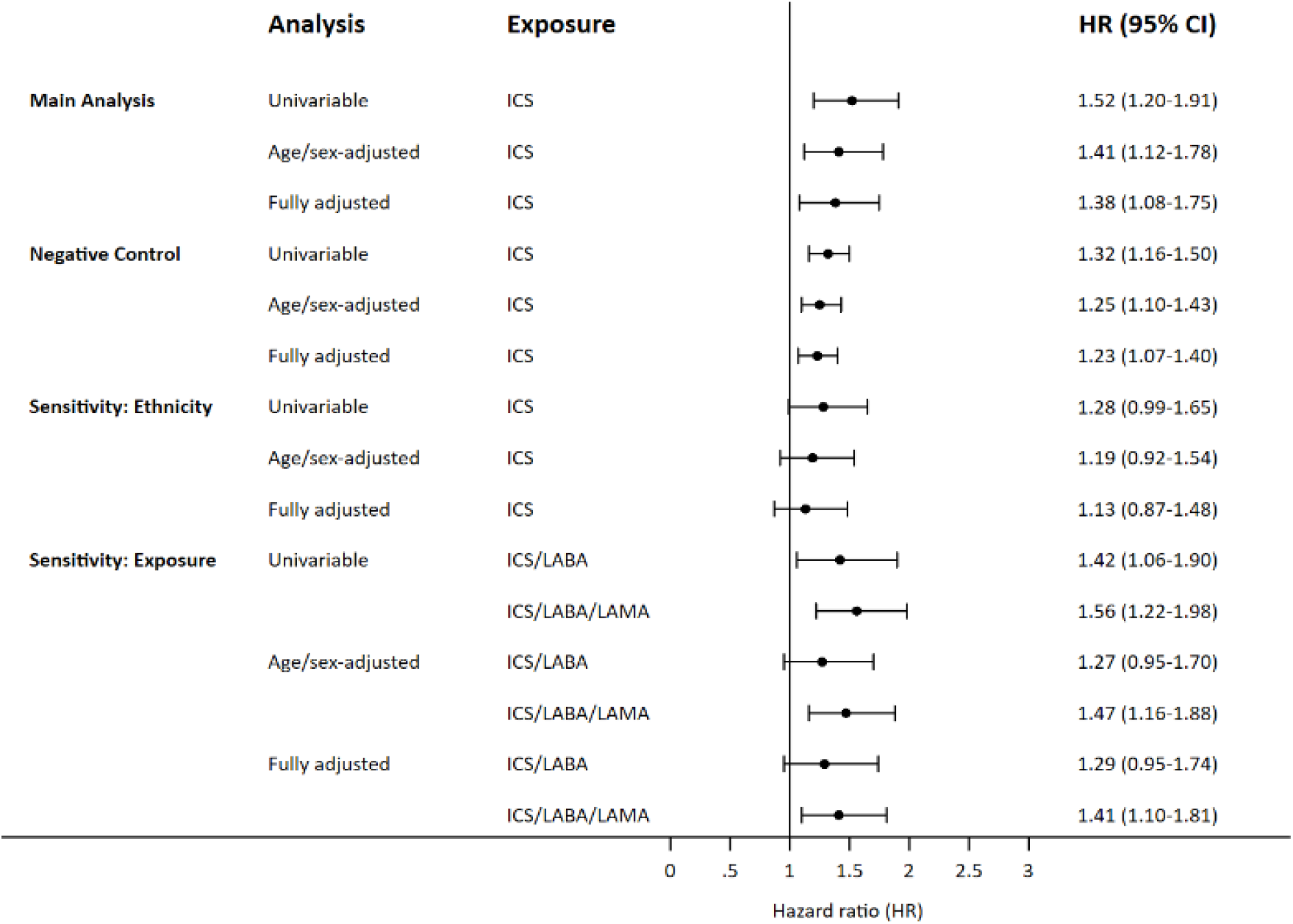
Univariable and Multivariable Models, COPD population.

#### Asthma

There were 515 COVID-19 related deaths in the treated asthma population; time to COVID-19 related death by treatment group can be seen in Figure 1b. In univariable models, receipt of both low/medium dose and high dose ICS was associated with an increased risk of COVID-19 related death (HR = 1.32, 95% CI = 0.98 - 1.77 and HR = 2.28, 1.62 - 3.20 respectively; Figure 3). These associations reduced markedly upon adjustment for age and gender, and the remaining pre-specified comorbidities (aHR = 1.10, 95% CI = 0.82 - 1.49 and aHR = 1.52, 95% CI = 1.08 - 2.14 for low/medium dose and high dose ICS respectively; Figure 3). Post-hoc analyses revealed that the greatest reduction in the strength of the association after the age and sex adjustment was adjustment for previous exacerbations (supplemental table 10). There was no evidence for a (pre-specified) interaction with age, and no deviations from the proportional hazards assumption (supplemental table 8 and 9, figure 7-12).

**Figure 3:**
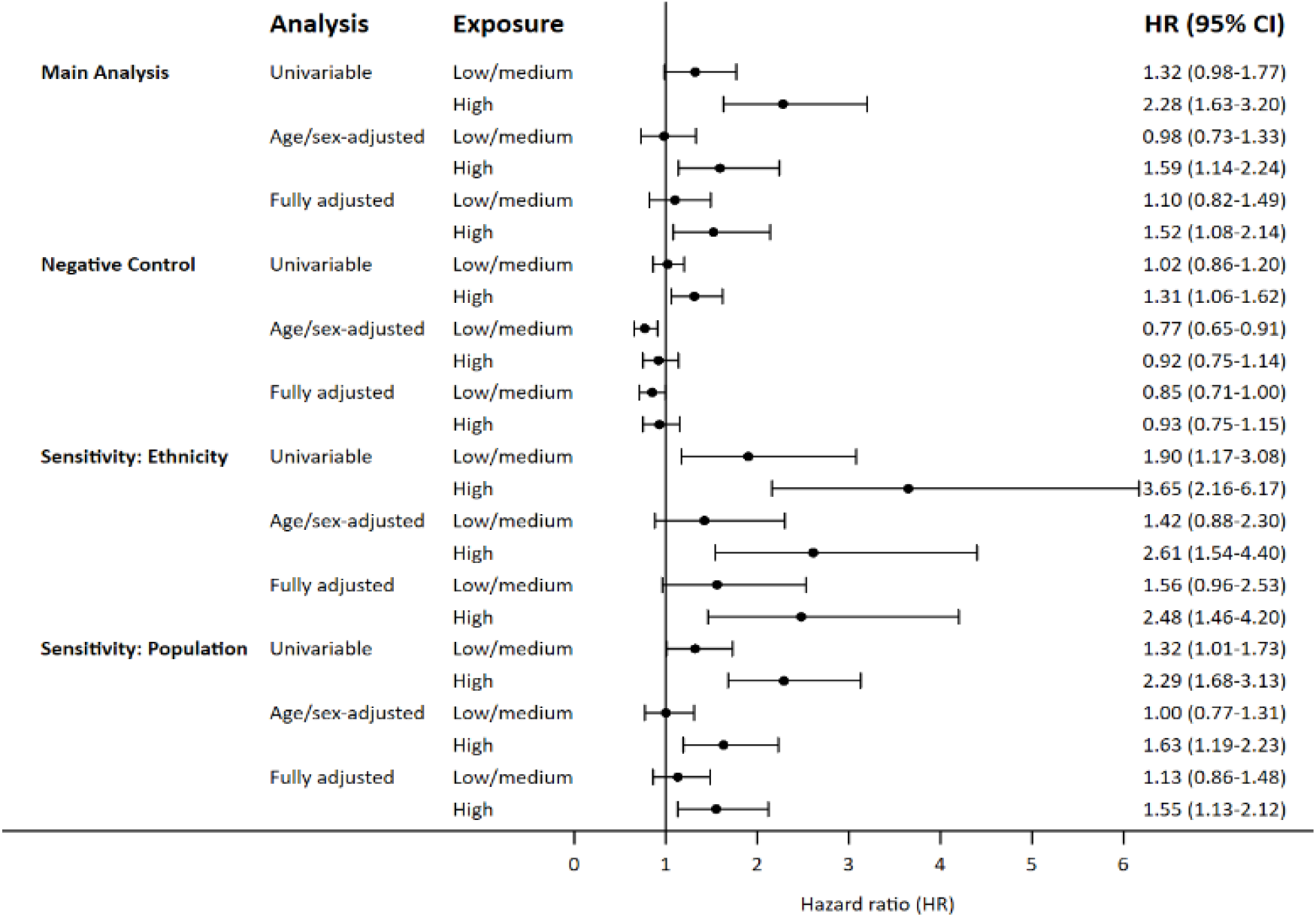
Univariable and Multivariable Models, Asthma population.

### Sensitivity Analyses

Figure 2 and 3 show results from sensitivity analyses. When considering receipt of ICS + LABA and ICS + LABA/LAMA separately in the COPD population, the risk of death was higher among those receiving ICS + LABA/LAMA (aHR = 1.41 95% CI = 1.10 - 1.81) but less markedly amongst those receiving ICS + LABA (aHR = 1.29, 95% CI = 0.95 - 1.74). Restricting analyses to people of white ethnicity to attempt to control for potential confounding by ethnicity led to a reduction in the hazard ratios in the COPD population (Figure 2), but not in the asthma population (Figure 3). Changing the population definition for the asthma population had a negligible impact on the results (Figure 3).

### Negative Control Analysis

In the COPD population, the risk of non-COVID-19 related death was higher among individuals in the ICS combination group with an adjusted HR = 1.23 (95% CI = 1.07 - 1.40). In the asthma population, there was no evidence of an association between receipt of high or low/medium dose ICS and non-COVID-19 related death with adjusted HRs of 0.85 (95% CI = 0.71 - 1.00) and 0.93 (95% CI = 0.75 - 1.15), respectively.

### Quantitative Bias Analysis

To fully explain the lower bound of the 95% CI in the COPD cohort (1.08), or for the high-dose ICS association in the asthma cohort (1.08), an unmeasured confounder would need to be associated (conditional on measured covariates) with either exposure or outcome by at least risk ratio (RR) 1.37 (e-value/high threshold) and with both exposure and outcome by at least RR 1.08 (low threshold; Figure 4a-b). An unmeasured confounder would need to have a stronger association with either exposure or outcome to move the observed HRs to a clinically meaningful protective effect of 0.8 (high threshold COPD: 2.84, asthma: 3.21, supplemental figure 14-15; low threshold COPD: 1.73, asthma: 1.90).

**Figure 4a:**
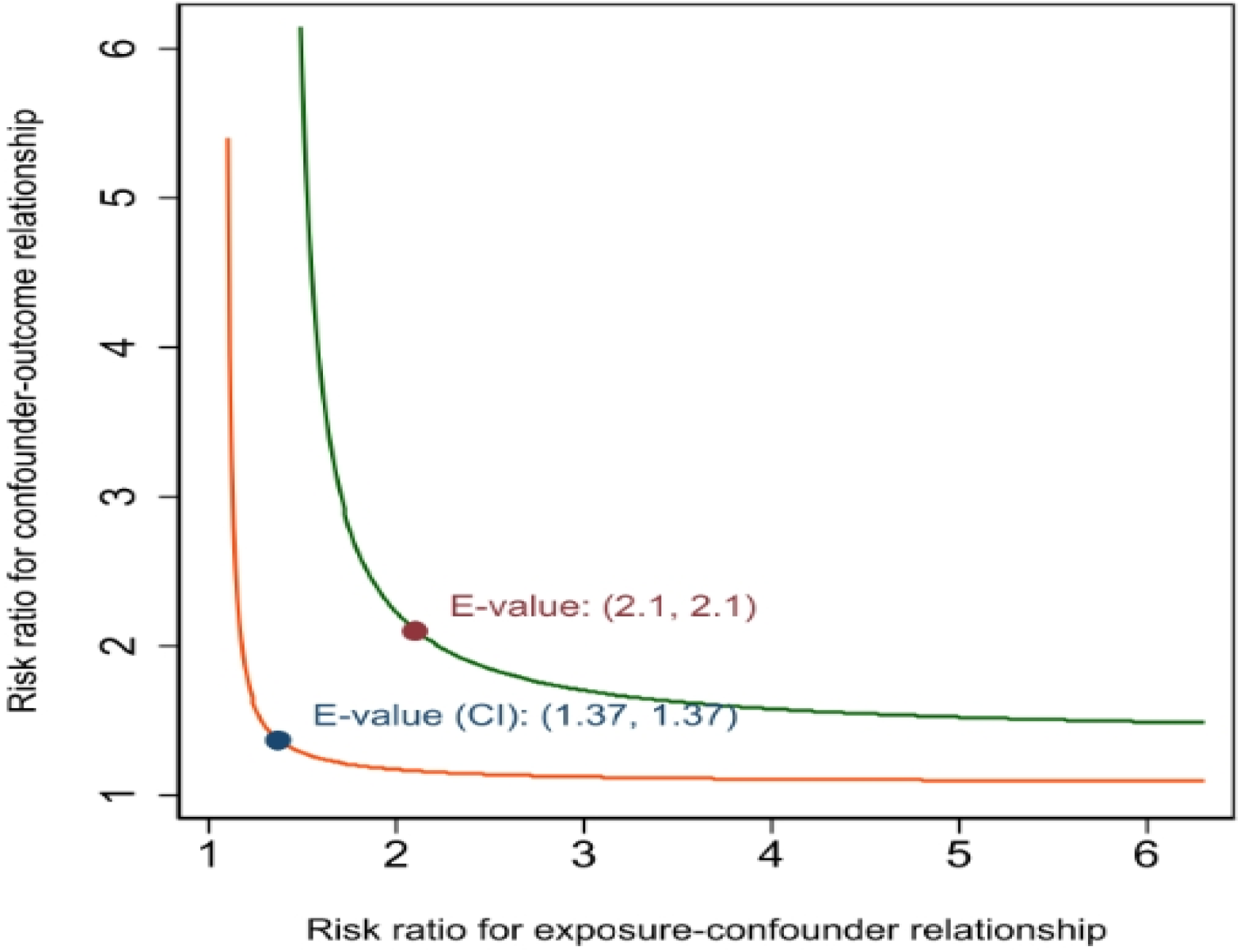
E-value for the lower 95% CI and point estimate in the COPD.

**Figure 4b:**
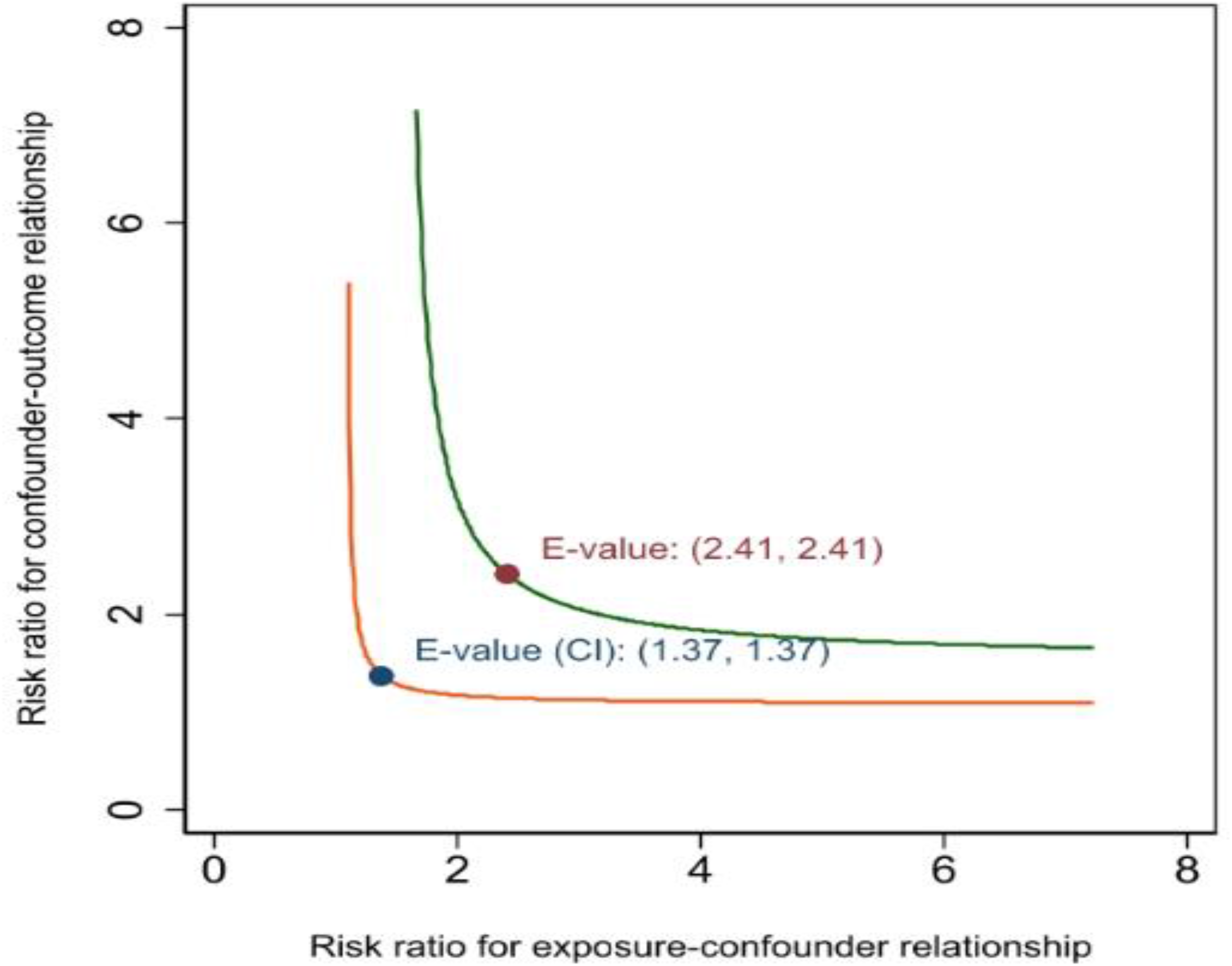
E-value for the lower 95% CI and point estimate in the Asthma population.

## Discussion

### Summary

This is the first study to investigate the association between regular ICS use and COVID-19 related death. Compared with non-ICS based treatments, ICS in people with COPD was associated with a ∼40% increased risk of COVID-19 related death. In people with asthma, high dose ICS use was associated with an ∼50% increased risk of COVID-19 related death, with little evidence of any association for low/medium dose ICS. Our findings do not provide any strong support for a protective effect from ICS use in these populations, as has been previously hypothesised. Our analyses overall indicate that the observed harmful associations could readily be explained by confounding due to underlying health differences between people prescribed ICS and those using other medications for asthma and COPD, rather than a causally harmful effect of ICS. Specifically, we observed a stronger association with COVID-19 related death between ICS triple therapy than ICS dual therapy in the COPD cohort; the ICS content of these two regimens is similar, and a causal effect of ICS would be expected to be comparable in these two groups. If we had successfully controlled for disease severity differences between treatment groups, we would also expect to see no association between ICS and the negative control outcome of non-COVID-19 death. The harmful association we observed suggests we had not perfectly captured all markers of disease severity, resulting in an association that is unlikely to be causal. The null finding between ICS and non-COVID-19 related death in the asthma cohort is less surprising, since asthma is less markedly associated with overall mortality compared to COPD^29^. Finally, quantitative bias analysis confirms that a hypothetical unmeasured confounder of modest strength could fully explain the observed results.

### Findings in Context

A literature review (box 2) found no other epidemiological studies or randomised controlled trials (RCTs) assessing the role of ICS in COVID-19. Two ongoing RCTs investigating the role of ICS in people hospitalised with laboratory confirmed COVID-19 and mild COVID-19 respectively (NCT04331054, NCT04330586) are due to complete later this year. The hypothesis of a protective effect of ICS in COVID-19 was based partly on the prevalence of chronic lung disease among outpatient and inpatient COVID-19 in China. However, more recent studies do not support initial assertions that people with chronic lung diseases (including COPD) are significantly underrepresented among COVID-19 patients ^6,7,30^. In addition, a number of studies have found that people with COPD are at greater risk of severe COVID-19 and death from COVID-19 once infected ^6,7^. The evidence for asthma in COVID-19 is more varied, with studies reporting both null and moderately harmful associations^6,7^. It may be that features other than ICS use, such as shielding, influence the risk of acquiring SARS-CoV-2 among asthmatics. Studies investigating the causal effect of chronic respiratory disease, including COPD, and asthma on SARS-CoV-2 infection risk and COVID-19 disease, ideally taking into account the relatively large degree of heterogeneity that exists within each of these diagnostic categories, are urgently needed to help inform about levels of risk for these patients.

### Strengths and weaknesses

The greatest strength of this study was the power we had to look at multiple drug treatments as our dataset included medical records from almost 24 million individuals. Our study is further strengthened by the use of two different study populations and active comparators, as well as sensitivity analyses to quantify the potential impact of unmeasured confounding on results. Another strength is our use of open methods: we pre-specified our analysis plan and have shared all analytical code.

We also recognise possible limitations. The primary limitation is the risk of confounding by indication due to unmeasured or imperfectly defined potential confounding variables. Decisions regarding treatment choices involve factors that may not be well recorded in electronic health records including measures such as spirometry, and likely steroid responsiveness. As we did not have secondary care data our assessment of exacerbation history was incomplete, limiting our ability to adjust for this. Our sensitivity analyses confirm that unmeasured confounding is a plausible reason for the harmful associations we observed. The proportional hazards assumption was not met for the COPD models, with KM plots indicating that the hazard ratio for this exposure increased over time. This is perhaps not surprising, as the risk of acquiring COVID-19 was lower in the early stages of the pandemic. The HR for the COPD population should be interpreted as an average over the entire follow-up period. Finally, it is important to note that the outcome of COVID-19 related death will reflect the risk both of becoming infected as well as the risk of developing severe disease and dying. It is possible that ICS use has a different effect on the risk of infection and on disease severity.

### Policy Implications and Future Research

We find no evidence that ICS has a strongly beneficial effect on COVID-19 related mortality, and therefore we cannot recommend that they are used to treat people with COVID-19 outside of the context of RCTs. Importantly, from the totality of the evidence provided here, including our sensitivity analyses, our results do not support the interpretation that regular ICS therapy for asthma or COPD increases risk of death from COVID-19, and do not provide evidence to support adjustments in ICS therapy among COVID-19 patients. Future observational studies of this clinical question are likely to face similar challenges around unmeasured confounding.

The UK has an unusually large volume of detailed longitudinal patient data. We have demonstrated that it is feasible to rapidly address specific hypotheses about medicines in a transparent manner inside the secure environment of an EHR vendor in order to minimise the large volumes of potentially disclosive data that would otherwise have to move into separate systems. We will use the OpenSAFELY platform to further inform the global response about drug treatments during the COVID-19 emergency.

### Summary

We found no evidence of a beneficial effect of regular ICS use on COVID-19 related mortality. Although we report a small harmful association, the pattern of results we observed suggests this could readily be explained by differences in underlying health between people receiving ICS and those receiving other respiratory medications. People currently taking ICS should continue taking them if recommended as part of routine care.

**Box 1: Prespecified hypothetical confounders**

- Age
- Sex
- Body Mass Index (BMI): Measurement of weight in the last decade
- Indices of multiple deprivation (IMD): Quintiles from IMD 2019
- Diagnosed hypertension
- Heart disease: categorised as heart failure and other heart disease
- Diabetes: categorised as controlled (HbA1c < 58 mmol/mol), uncontrolled (HbA1c
- ≥ 58 mmol/mol) or HbA1c not measured measured within the last 12 months
- Cancer
- Immunosuppressive conditions: organ transplant, sickle cell anaemia and splenectomy
- Chronic kidney disease: based on creatinine measurements within the last 12 months or ever having a code for renal dialysis
- Influenza vaccination status: recorded between 01Sep 2019 and 01 Mar 2020
- Pneumococcal vaccination status: record in the five years prior to 01 Mar 2020
- Statin use: recorded within four months prior to 1st of March 2020,
- Exacerbation history: different methods used for asthma ^31^ and COPD population ^32^with COPD models additionally adjusted for a history of asthma.

**Box 2: Research in Context**

**Evidence before this study**

At the start of the global coronavirus outbreak, Inhaled Corticosteroids (ICS) were hypothesised to offer some protection against either infection with SARS-CoV-2 or against severe outcomes from COVID-19, such as acute respiratory distress syndrome (ARDS) and respiratory failure^9^, despite these medications being known to increase the risk of pneumonia and other respiratory tract infections^17,33^. The hypothesis was based at least in part on epidemiological data showing a low prevalence of chronic respiratory disease among Chinese COVID-19 patients^2^, although there was also some support of a potential protective effect from in-vitro studies^12,13^. Most recently, ICS exposure has been found to correlate with a lower expression of ACE2 and TMPRSS2, the entry receptors used by SARS-CoV-2, in sputum cells^34^. A recent systematic review evaluating whether administration of ICS was associated with clinical outcomes in COVID-19, SARS or MERS identified no relevant studies^10^.

**Added value of this study**

Our study was specifically designed to assess the role of pre-morbid ICS use in COVID-19. We included two cohorts of participants: people with asthma, and people with COPD, both of whom have a possible indication for ICS. Neither analysis was strongly suggestive that regular ICS therapy for asthma or COPD has a clinically important causal effect on COVID-19 mortality in either direction.

Our study has several key strengths: Firstly, it includes almost a million participants making it the largest contemporary study of ICS use to date. Secondly, we used active comparators and multiple sensitivity analyses to reduce and quantify the impact of possible unmeasured confounding. Finally, our analyses were pre-specified and we used open methods throughout the study with code and codelists available for examination and reuse.

**Implications of all the available evidence**

Evidence from our study and other research suggests there is neither a demonstrable benefit nor clear harm from ICS use against COVID-19 related mortality and therefore at present there is no evidence people should alter their ICS therapies during the pandemic. We also cannot recommend that ICS be used specifically to treat people with COVID-19 outside of the context of clinical trials. Future observational research is likely to be subject to similar issues around unmeasured confounding, and evidence from RCTs will provide more definitive answers regarding the role of ICS in COVID-19 later this year.

## Administrative

## Acknowledgements

We are very grateful for all the support received from the TPP Technical Operations team throughout this work; for generous assistance from the information governance and database teams at NHS England / NHSX.

## Ethical approval

This study was approved by the Health Research Authority (REC reference 20/LO/0651) and by the LSHTM Ethics Board (ref 21863). No further ethical or research governance approval was required by the University of Oxford but copies of the approval documents were reviewed and held on record.

## Guarantor

BG

## Contributorship

Contributions are as follows:

Conceptualization LS BG ID;

Data curation CB JP JC SH SB DE PI CM;

Formal Analysis AS AJW BM CM JB;

Funding acquisition BG LS;

Information governance AM BG CB JP;

Methodology ID AS LT CTR AW KB EW SJWE JQ LS JB CM AJW BM SB BG;

Disease category conceptualisation and codelists CM AJW AS CTR PI SB DE CB JC JP SH HD HC KB SB AM BM LT ID HM RM HF JQ;

Ethics approval HC EW LS BG;

Project administration AS CM HC CB SB AM LS BG; Resources BG LS;

Software SB DE PI AJW CM CB FH JC SH;

Supervision ID LS BG; Visualisation CTR AJW;

Writing (original draft) AS ID CM;

Writing (review & editing) AS AJW BM CM CTR KB EW HJC HD SB CB AM DE PI HM LT RH KW AYSW HF RC SJWE JQ LS ID BG;

All authors were involved in design and conceptual development and reviewed and approved the final manuscript.

